# Ten more years of maternity claims: what can another decade of NHS litigation data tell us about mode of birth, informed consent and patient safety?

**DOI:** 10.64898/2026.05.01.26352218

**Authors:** Jonathan H West, Myles J Taylor, Michael Magro

**Affiliations:** Former NHS Consultant in Obstetrics and Gynaecology (FRCS, FRCOG), Exeter, UK; Royal Devon University Healthcare NHS Foundation Trust (Royal Devon and Exeter Hospital), Exeter, UK; Barking, Havering and Redbridge University Hospitals NHS Trust, London, UK; former Darzi Fellow, NHS Resolution (2016–17)

**Author notes:** Correspondence: Jonathan H West, Exeter, UK.

**Keywords:** caesarean section, clinical negligence, health economics, informed consent, litigation, mode of birth

## Abstract

**Objective:** To quantify the contribution of harm arising in labour to NHS obstetric litigation costs, compare the pathway costs of planned vaginal birth (PVB) and planned caesarean section (PCS), and review consent for planned mode of birth.

**Design:** Secondary analysis of administrative litigation and cost data, with policy review.

**Setting:** NHS England, 2015/16 to 2024/25.

**Population:** 11,881 obstetric claims notified to NHS Resolution; national NHS delivery activity.

**Methods:** Freedom of Information data on claims by primary cause and injury. Activity-weighted National Schedule of NHS Costs reference costs for 2023/24 and 2024/25. Indemnity attributed to labour by cause code (primary analysis) and by injury (sensitivity analysis). Consent guidance reviewed against *Montgomery v Lanarkshire Health Board*.

**Main outcome measures:** Damages and total payments by cause and injury; per-birth cost difference between PVB and PCS pathways.

**Results:** 7,216 claims were settled, with total payments of £5,974 million (damages £4,971 million). Eleven labour-exclusive cause codes accounted for 46.5% of total payments; cerebral palsy and brain damage accounted for 73.3%. In 2024/25, reference costs were £6,202 per PCS and £6,337 per PVB pathway. Including labour-attributable indemnity, the PVB pathway exceeded PCS by £879–£2,582 per birth (cause-code basis, cash to actuarial) and £1,190–£4,142 under injury-based sensitivity analyses. No formal comparative-risk consent process exists for women planning vaginal birth.

**Conclusions:** Obstetric litigation costs are dominated by harm arising in labour. On current NHS cost data, PVB is not the cheaper pathway. Comparative-risk counselling on planned mode of birth should be offered to all women at booking.

**Tweetable abstract:** Labour harm dominates NHS maternity claims; planned vaginal birth is not cheaper than planned caesarean.

## Introduction

In October 2012 the NHS Litigation Authority (NHSLA, now NHS Resolution) published *Ten Years of Maternity Claims*, covering claims arising between 2000 and 2010.^1^ It identified 5,087 claims totalling approximately £3.12 billion. Its cause-code data indicated that management of labour and cerebral palsy were the dominant cost categories,^2^ a finding this paper extends and quantifies.

The view that planned caesarean section (PCS) costs the NHS more than planned vaginal birth (PVB) has long influenced policy. NICE Clinical Guideline CG132 (2011) concluded that PVB was approximately £700 cheaper than a maternal-request caesarean, and estimated savings of £4.9 million for every percentage-point reduction in the caesarean rate.^3^ That analysis predated any systematic attribution of indemnity costs to mode of birth. It acknowledged that PCS might be less costly once downstream costs were included, but this was not carried into its headline conclusions.

Since then, the Ockenden review,^4^ the East Kent investigation,^5^ Each Baby Counts,^6^ the Maternity Incentive Scheme and the Early Notification Scheme have all targeted intrapartum harm. Both investigations described cultures in which normal birth was promoted as an ideal and women had limited freedom to choose PCS, contributing to preventable harm.^4, 5^ Meanwhile, *Montgomery v Lanarkshire Health Board* established that patients must be told of material risks and reasonable alternatives.^7^

This study aimed to: (i) quantify the contribution of harm arising in labour to obstetric indemnity costs, using ten years of NHS Resolution claims data; (ii) compare pathway-level NHS reference costs of PVB and PCS across two consecutive years; and (iii) examine the consent architecture for planned mode of birth.

## Methods

All financial values are nominal unless stated. “Planned caesarean section” and “planned caesarean birth” are used synonymously.

### Claims data

Obstetric claims data for 2015/16 to 2024/25 were obtained from NHS Resolution under the Freedom of Information Act 2000 (FOI_7787), broken down by year, primary cause and primary injury.^8^ Supplementary data on claims with cerebral palsy or brain damage (CP/BD) as the primary injury for 2019/20 to 2024/25 came from FOI_7633,^9^ and annual obstetric totals for the same period from FOI_7770.^10^

NHS Resolution reports, for each claim closed with damages, the damages paid, NHS legal costs and claimant legal costs. “Total paid” is the sum of all three. Each is paid to the end of the year of closure, or of settlement for periodical payment orders. Proportions are calculated on a consistent basis throughout, numerator and denominator both on total paid unless stated otherwise.

### Labour attribution: primary analysis by cause code

NHS Resolution records a single primary cause per claim. Eleven cause codes were classed as labour-exclusive. Four are intrapartum monitoring failures: “Fail To Make Resp To Abnrm FHR”, “Fail To Monitor 2nd Stg Labour”, “Fail To Monitor 1st Stg Labour” and “Fail Mon Dose/rate Syntocinon”. Seven are complications that occur only in labour or vaginal delivery: “Perineal Tear-1st,2nd,3rd Deg”, “Labial Tear”, “Forceps Delivery”, “Inapp Use Of Forceps/Ventouse”, “Application Of Excess Force”, “Fail To Correctly Apply Forcep” and “Repeat Attempt Forcep/Ventouse”. The FHR response code could in principle include antepartum CTG events, but a thematic review of 50 NHS Resolution cerebral palsy claims found that all but one (98%) were directly attributable to intrapartum causes.^11^

Intrapartum harm recorded under generic codes such as “Fail / Delay Treatment” or “Fail To Recog. Complication Of” is excluded. The cause-code attribution is therefore a lower bound.

### Labour attribution: sensitivity analysis by injury

No cross-tabulation of cause by injury was available. As a sensitivity analysis, indemnity for claims with cerebral palsy or brain damage as the primary injury was attributed to labour under three assumptions: 90%, 98% (the proportion found in the thematic review^11^) and 100% intrapartum origin. A further upper scenario added Erb’s palsy, which arises almost exclusively at vaginal delivery.

### Pathway costs

Activity and unit costs by Healthcare Resource Group (HRG) were taken from the NHS England National Schedule of NHS Costs for 2024/25^12^ and 2023/24.^13^ These are reported reference costs, not Payment Scheme prices. The PVB pathway cost was the activity-weighted average of normal vaginal delivery (NZ30–34), assisted vaginal delivery (NZ40– 44) and emergency caesarean section (NZ51). The PCS cost was the activity-weighted average of NZ50A–C. For 2023/24, national averages were rebuilt from the provider-level source file (Organisation level source data part 1), summing cost and activity across all providers and settings; activity counts under 8, suppressed in that file, were estimated from each row’s cost and national mean. The maternity-specific CNST uplift in the 2024/25 NHS Payment Scheme is applied uniformly across maternity HRGs.^14^

### Indemnity per birth

Labour-attributable indemnity per birth was calculated on two bases: the 2024/25 obstetric total paid of £760 million (cash basis)^10^ and NHS Resolution’s 2024/25 actuarial obstetric cost of harm of £2,500 million.^15^ The labour-exposed denominator of 474,828 births is 591,759 total deliveries in 2019/20^16^ less the 19.76% planned caesarean fraction. All labour-attributable indemnity was assigned to the PVB pathway and added to the reference-cost differential.

### Consent architecture

Consent and guidance for planned mode of birth were reviewed against RCOG Consent Advice No. 14,^17^ NICE NG192^18^ and *Montgomery*.^7^

## Results

### Volume and cost of obstetric claims

Between 2015/16 and 2024/25, 11,881 obstetric claims were notified, approximately one per 500 NHS births in England.^8^ Of these, 7,216 closed with damages, with total payments of £5,974 million: damages £4,971 million, NHS legal costs £233 million and claimant legal costs £770 million. A further 4,944 claims closed with nil damages, incurring £41 million in legal costs.

Annual total payments rose from £321 million in 2015/16 to a peak of £771 million in 2023/24, and were £760 million in 2024/25 (damages £626 million), a 2.4-fold nominal increase over the decade.^8, 10^ Annual CP/BD payments are shown in Table 3.

### Primary causes and labour attribution

The eleven labour-exclusive cause codes accounted for 1,733 settled claims and £2,776 million in total payments, 46.5% of all obstetric total payments (Table 2). On a damages- only basis the share was 47.7% (£2,373 million of £4,971 million). The four intrapartum monitoring codes alone accounted for £2,491 million.

Three generic codes that include intrapartum events, “Fail / Delay Treatment” (£725 million), “Fail To Recog. Complication Of” (£432 million) and “Failure/Delay Diagnosis” (£223 million), together accounted for a further £1,380 million (23.1%). These cannot be divided by timing in the available data.

“Fail To Warn – Informed Consent” accounted for 197 settled claims and £153 million in total payments (damages £128 million), with 461 claims notified. This probably understates consent-related liability, because inadequate consent is often a contributing rather than primary cause.

### Cerebral palsy and brain damage

Over the ten years, claims with cerebral palsy (685) or brain damage (441) as the primary injury made up 15.6% of settled claims but £4,376 million, 73.3% of total payments (77.7% of damages).^8^ The six-year data from FOI_7633 give the same concentration: 16.6% of settled claims by volume and 77.7% of damages, at a mean of £3.58 million per claim, rising to 84.0% of damages in 2024/25 (Table 3).^9, 10^ Erb’s palsy accounted for a further £170 million (2.8%).

### Pathway reference costs

In 2024/25 the activity-weighted reference cost of PCS was £6,202 per birth, £135 less than the PVB pathway at £6,337 (Table 1).^12^ In 2023/24, rebuilt from provider-level source data, the corresponding costs were approximately £5,790 for PCS and £5,720 for PVB, with PVB about £70 less costly.^13^ The two pathways were therefore at near parity in both years, within about 2% of each other. Between the two years, unit costs rose by approximately 7% (PCS) and 11% (PVB).

**Table 1.** Activity-weighted reference costs for the planned vaginal birth (PVB) and planned caesarean section (PCS) pathways, England 2024/25.

| HRG | Description | Episodes | Share of PVB pathway | Weighted unit cost |
| --- | --- | --- | --- | --- |
| NZ30–34 | Normal vaginal delivery | 236,674 | 55.2% | £4,733 |
| NZ40–44 | Assisted vaginal delivery | 59,608 | 13.9% | £6,925 |
| NZ51 | Emergency caesarean section | 132,136 | 30.8% | £8,945 |
| PVB pathway | NZ30–34 + NZ40–44 + NZ51 | 428,418 | 100% | £6,337 |
| NZ50 | Planned caesarean section | 105,502 | — | £6,202 |
| Difference | PVB minus PCS |  |  | £135 |

Planned caesarean rate: NZ50 as a share of all 533,920 delivery episodes = 19.8%. Source: National Schedule of NHS Costs 2024/25, admitted patient care, national activity and unit cost columns, all settings.<sup>12</sup> 2023/24 for comparison, rebuilt from Organisation level source data part 1 (all providers and settings; suppressed activity estimated): NZ50 approximately 97,300 episodes at £5,790; PVB pathway approximately 435,300 episodes at £5,720.<sup>13</sup>

**Table 2.** Labour-exclusive cause codes, obstetric claims closed with damages, 2015/16– 2024/25.

| Cause code | Claims | Damages (£m) | Total paid (£m) | Mean total paid per claim (£m) |
| --- | --- | --- | --- | --- |
| *Intrapartum monitoring failure* |  |  |  |  |
| Fail To Make Resp To Abnrm FHR | 418 | 936.2 | 1,069.3 | 2.56 |
| Fail To Monitor 2nd Stg Labour | 477 | 792.7 | 914.4 | 1.92 |
| Fail To Monitor 1st Stg Labour | 269 | 365.5 | 427.1 | 1.59 |
| Fail Mon Dose/rate Syntocinon | 33 | 70.4 | 80.5 | 2.44 |
| *Labour-exclusive delivery complications* |  |  |  |  |
| Application Of Excess Force | 106 | 58.9 | 78.3 | 0.74 |
| Forceps Delivery | 97 | 49.7 | 65.4 | 0.67 |
| Perineal Tear-1st,2nd,3rd Deg | 168 | 35.5 | 56.4 | 0.34 |
| Inapp Use Of Forceps/Ventouse | 73 | 33.2 | 43.1 | 0.59 |
| Fail To Correctly Apply Forcep | 50 | 25.0 | 31.4 | 0.63 |
| Repeat Attempt Forcep/Ventouse | 13 | 4.3 | 6.3 | 0.48 |
| Labial Tear | 29 | 1.8 | 4.0 | 0.14 |
| **Eleven codes combined** | **1,733** | **2,373.2** | **2,776.1** |  |
| **Share of all obstetric claims** | 24.0% | **47.7%** | **46.5%** |  |
| *Selected other causes* |  |  |  |  |
| Fail / Delay Treatment | 1,336 | 585.2 | 725.5 | 0.54 |
| Fail To Recog. Complication Of | 491 | 357.9 | 431.6 | 0.88 |
| Fail Antenatal Screening | 325 | 314.3 | 368.8 | 1.13 |
| Failure/Delay Diagnosis | 619 | 161.8 | 223.2 | 0.36 |
| Birth Defects | 74 | 182.7 | 205.3 | 2.77 |
| Fail To Warn-Informed Consent | 197 | 128.0 | 153.0 | 0.78 |
| **All obstetric claims** | **7,216** | **4,970.6** | **5,974.2** | 0.83 |

Total paid = damages + NHS legal costs + claimant legal costs. Source: FOI\_7787, Tables 4 and 5.<sup>8</sup>

**Table 3.** Cerebral palsy and brain damage (CP/BD) claims as a proportion of obstetric damages, 2019/20–2024/25.

| Year | CP/BD claims | CP/BD damages (£m) | All obstetric damages (£m) | CP/BD share of damages |
| --- | --- | --- | --- | --- |
| 2019/20 | 113 | 397 | 495 | 80.2% |
| 2020/21 | 111 | 389 | 502 | 77.5% |
| 2021/22 | 106 | 354 | 504 | 70.2% |
| 2022/23 | 132 | 473 | 597 | 79.2% |
| 2023/24 | 124 | 486 | 655 | 74.2% |
| 2024/25 | 147 | 526 | 626 | 84.0% |
| **Total** | **733** | **2,625** | **3,378** | **77.7%** |

CP/BD = cerebral palsy or brain damage as primary injury. 733 of 4,416 settled claims (16.6%); mean damages £3.58m per CP/BD claim versus £0.77m for all obstetric claims. A further 604 CP/BD claims closed with nil damages (£13.4m legal costs). Damages only; legal costs excluded. Sources: FOI\_7633<sup>9</sup> and FOI\_7770.<sup>10</sup>

### Indemnity per birth

Applying the 46.5% cause-code share to the 2024/25 cash total of £760 million gives £353 million of labour-attributable indemnity, or £743 per labour-exposed birth (Table 4). On the actuarial basis, NHS Resolution’s 2024/25 obstetric cost of harm was £2,500 million, 54.5% of the whole NHS clinical negligence actuarial cost;^15^ the same share gives £1,162 million, or £2,447 per labour-exposed birth. Adding the £135 reference-cost differential, the PVB pathway exceeded PCS by **£879 per birth (cash) to £2**,**582 (actuarial)**.

**Table 4.**
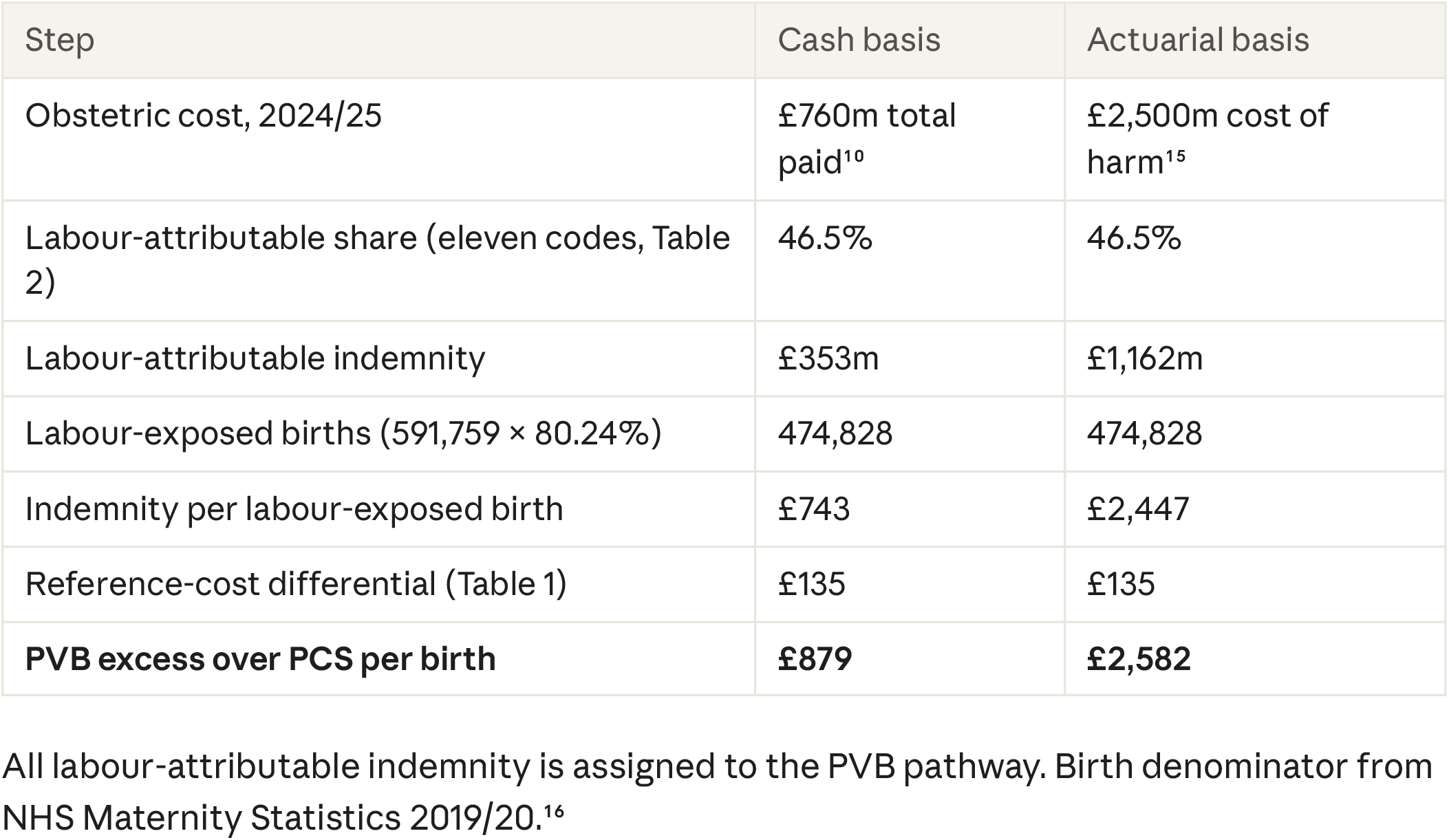
Labour-attributable indemnity and the PVB excess over PCS: primary (cause- code) analysis.

Under the injury-based sensitivity analysis the excess rises to £1,190–£3,606 (CP/BD 90% intrapartum), £1,284–£3,915 (98%, as observed in the thematic review) and £1,307–£3,992 (100%), and to £1,353–£4,142 with Erb’s palsy added (Table 5). Every alternative attribution increases the excess.

**Table 5.** Sensitivity analysis: PVB excess over PCS per birth under alternative labour attributions.

| Attribution | Labour share of total paid | Cash basis | Actuarial basis |
| --- | --- | --- | --- |
| Cause code (primary, lower bound) | 46.5% | £879 | £2,582 |
| CP/BD, 90% intrapartum | 65.9% | £1,190 | £3,606 |
| CP/BD, 98% intrapartum (thematic review<sup>11</sup>) | 71.8% | £1,284 | £3,915 |
| CP/BD, 100% intrapartum | 73.3% | £1,307 | £3,992 |
| CP/BD 100% plus Erb's palsy | 76.1% | £1,353 | £4,142 |

### Consent architecture

RCOG Consent Advice No. 14 provides a formal consent form for planned caesarean birth, including a comparative risk table covering both modes of birth, with a clinician’s signature confirming that the risks of the alternatives were discussed.^17^ No equivalent document exists for PVB. NICE NG192 requires a comparative risk discussion only when a woman requests PCS.^18^ There is no formal process at booking for all women to be counselled on planned mode of birth, no comparative risk table and no requirement to document the discussion (Appendix Table A1).

## Discussion

### Main findings

Harm arising in labour remains the main driver of NHS obstetric litigation. Labour- exclusive cause codes account for at least 46.5% of a decade’s £6.0 billion in payments, and cerebral palsy and brain damage, overwhelmingly intrapartum in origin, for 73.3%. On NHS reference costs, the two pathways were at near parity in both 2023/24 and 2024/25. Including labour-attributable indemnity, PVB exceeds PCS by £879–£2,582 per birth on the conservative attribution, and by more under every alternative. No formal comparative-risk consent process exists for women planning vaginal birth.

### Strengths and limitations

The study uses complete national claims data for a decade, obtained under FOI and publicly available, and reports both cash and actuarial bases. The primary attribution is deliberately conservative, and the sensitivity analysis shows the direction of the finding is robust to the attribution method.

NHS Resolution records a single primary cause and injury per claim, with no coded field for timing (antenatal, intrapartum, postnatal) or procedure.^8^ The cause-code attribution excludes intrapartum harm recorded under generic codes, and without a cause-by-injury cross-tabulation the injury-based estimates rest on stated assumptions. Conversely, a small amount of indemnity arising from caesarean operations themselves (for example intra-operative injury or retained swabs) was not assigned to the PCS pathway; these codes total under 2% of payments and some arise from emergency caesareans within the PVB pathway.

Claims data reflect year of closure, not incident year, and exclude periodical-payment damages due after the settlement year, so cash figures understate eventual liability. Some inputs are approximate: suppressed activity counts in the 2023/24 source data were estimated, and a 2019/20 birth denominator is combined with 2024/25 indemnity. These approximations do not change the direction or order of magnitude of the results.

Reference costs are not a full economic comparison: downstream costs of caesarean in later pregnancies, such as placenta accreta spectrum and repeat surgery, are not captured. On NHS Payment Scheme delivery-phase prices, which are modelled from older cost data and sit well below reported costs, planned caesarean is approximately £200 more than the PVB pathway; including labour-attributable indemnity reverses this.^14^ Compensated harm also does not capture all avoidable harm. The consent analysis is not legal advice.

### Interpretation

#### The persistence of intrapartum harm

Annual obstetric payments more than doubled in nominal terms over the decade despite unprecedented national attention to maternity safety. The concentration of cost in labour-related mechanisms and in cerebral palsy and brain damage has been stable across datasets spanning a quarter of a century,^1, 11^ a pattern consistently associated with failures of CTG interpretation and response. This suggests current approaches are not reaching the underlying causal mechanisms. Neither the NHS Resolution database nor the Maternity Services Data Set^19^ supports causal analysis at population level, so national intervention remains necessarily approximate.

#### The economics

The common presumption that PVB is the cheaper pathway is not supported by current NHS data. NICE’s position that litigation costs should be excluded from economic models^20^ sits uneasily with costs that NHS trusts actually bear through CNST contributions, a tension noted in relation to CG132. ^21^ The maternity CNST uplift is also applied uniformly across all delivery HRGs,^14^ although the cause-code evidence shows liability is concentrated in labour-dependent mechanisms. This allocation should be reviewed, and NICE’s economic analysis of mode of birth updated to include indemnity.

#### The consent gap and *Montgomery*

“Fail To Warn – Informed Consent” generated £153 million in payments, with 461 claims notified, reflecting the post-*Montgomery* landscape.^7, 22^ The asymmetry in consent has three consequences. Medicolegally, any case of intrapartum harm without documented comparative-risk counselling carries potential *Montgomery* exposure. For policy, the absence of documentation prevents study of the relationship between consent quality and outcomes. Ethically, women choosing the pathway that generates most litigation receive less formal information than those choosing PCS.

#### Operational context as material information

Clinicians reassess planned mode of birth throughout pregnancy and labour. Under *Montgomery*, the test is what a reasonable person in the patient’s position would regard as relevant.^7, 23^ Delays in scheduling induction at times of high demand, uncertain out-of-hours anaesthetic or senior obstetric cover, and reliance on locum staff are circumstances a reasonable woman might regard as material. A full discussion on admission in active labour would be inappropriate. A practical alternative is documented counselling at booking that planned mode of birth may be revisited in the light of clinical and operational circumstances on admission.

#### The NICE counselling pathway

NICE NG192 recommendations 1.2.26–1.2.31 support a woman’s choice of PCS after informed discussion, and recommendation 1.2.30 is clear that her decision should be respected.^18^ The problem is architectural rather than attitudinal: the comparative-risk discussion (1.2.27) is triggered only by a request for PCS. PCS is treated as the departure requiring explanation and PVB as the default requiring none.

Anchoring the comparative-risk discussion at booking for all women would correct this, while retaining psychological referral for tokophobia. Women would then receive what those choosing PCS already receive under Consent Advice No. 14,^17^ with a free choice recorded in the notes.

## Conclusion

The individual probability of a serious adverse event in childbirth leading to a claim is low, but risk-benefit evaluation must also weigh the severity of harm and the pathway through which it arises.^23^ PCS and PVB have different profiles of complications, and a woman may make a rational choice between them; she should be properly counselled to do so.

Three structural changes follow. First, the booking appointment should be a formal comparative-risk consent point for all women, equivalent to the process currently required for PCS under Consent Advice No. 14. Second, a documentation framework should be developed for disclosing operational and resource factors that materially affect intrapartum safety. Third, CNST costs should be allocated in proportion to the pathway-level mechanisms that generate them, and NICE’s economic analysis of mode of birth should include indemnity.

The persistent failure of clinical programmes to reduce intrapartum harm suggests that correcting the consent architecture, which operates through informed patient choice rather than further system-level clinical change, may be a tractable approach. A further priority is a data architecture able to represent clinical trajectories in confidentiality- preserving form, so that causal analysis becomes possible at population level.

## Data Availability

Data availability: The NHS Resolution FOI responses, NHS England HRG cost extract, and derived analysis files supporting this study are openly available in Zenodo at [https://doi.org/10.5281/zenodo.19788703]. Original FOI responses are also available directly from NHS Resolution at the URLs cited in the reference list. All other principal sources are in the public domain.

https://resolution.nhs.uk/wp-content/uploads/2026/02/FOI_7633_Claims-under-ELSGP-and-Covid-scheme.pdf

https://resolution.nhs.uk/wp-content/uploads/2026/04/FOI_7770_Clinical-Claims-under-specialty-Obstetrics.pdf

https://assets.publishing.service.gov.uk/media/687a2dae312ee8a5f0806b6f/nhs-resolution-annual-report-and-accounts-2024-to-2025-hc1065-web-accessible.pdf

https://www.england.nhs.uk/wp-content/uploads/2025/11/NCC_National-Schedule_2024_25.zip

https://www.england.nhs.uk/publication/2023-24-national-cost-collection-data-publication/

https://www.england.nhs.uk/publication/2023-25-nhs-payment-scheme/

https://digital.nhs.uk/data-and-information/publications/statistical/nhs-maternity-statistics

## Declarations

### Contribution to authorship

JHW conceived the study, obtained the FOI data, performed the analysis and drafted the manuscript. MJT and MM contributed to interpretation and critically revised the manuscript. All authors approved the final version.

### Details of ethics approval

Not required. The study analyses aggregated, anonymised administrative data released under the Freedom of Information Act 2000 and published sources.

### Patient and public involvement

No patients or members of the public were involved in the design or conduct of this study.

### Data availability

Supporting data and a formula-driven calculation workbook are deposited on Zenodo (doi:10.5281/zenodo.19788703; version 2 corrects version 1). The NHS Resolution FOI responses and NHS England cost data are publicly available at the URLs cited.

### Version history

This is version 3 of this preprint. It corrects errors in earlier versions: the labour-attributable share of obstetric litigation payments (46.5%, previously reported as 55.9% through mixing damages and total-paid figures) and the 2023/24 comparison reference costs (previously day-case values rather than all-settings averages). All dependent figures have been recalculated. The conclusions are unchanged.

---

**Appendix Table A1.** Consent architecture for planned mode of birth.

| Feature | Planned caesarean birth | Planned vaginal birth |
| --- | --- | --- |
| Formal consent document | RCOG Consent Advice No. 14<sup>17</sup> | None |
| Comparative risk table | Yes, in consent form | No |
| Timing of consent process | Antenatal, before planned procedure | No formal process at booking or later |
| Documentation of risk discussion | Clinician signs that risks of both modes were discussed | No equivalent requirement |
| *Montgomery* compliance<sup>7</sup> | Partly addressed: comparative risks of PVB presented | Structural gap: no formal comparative-risk counselling |
| Operational and resource disclosure | Not applicable | No framework for disclosing staffing, theatre availability or decision-to-delivery performance |
| NICE pathway<sup>18</sup> | Not applicable | Counselling triggered only by a PCS request (NG192 1.2.26–1.2.30); none triggered by choosing PVB |

Sources: RCOG Consent Advice No. 14;<sup>17</sup> NICE NG192;<sup>18</sup> *Montgomery*;<sup>7</sup> GMC Decision Making and Consent.<sup>23</sup>

